# Modelling and analysis of COVID-19 epidemic in India

**DOI:** 10.1101/2020.04.12.20062794

**Authors:** Alok Tiwari

## Abstract

COVID-19 epidemic is declared as the public health emergency of international concern by the World Health Organisation in the second week of March 2020. This disease originated from China in December 2019 has already caused havoc around the world, including India. The first case in India was reported on 30^th^ January 2020, with the cases crossing 6000 on the day paper was written. Complete lockdown of the nation for 21 days and immediate isolation of infected cases are the proactive steps taken by the authorities. For a better understanding of the evolution of COVID-19 in the country, Susceptible-Infectious-Quarantined-Recovered (SIQR) model is used in this paper. It is predicted that actual infectious population is ten times the reported positive case (quarantined) in the country. Also, a single case can infect 1.55 more individuals of the population. Epidemic doubling time is estimated to be around 4.1 days. All indicators are compared with Brazil and Italy as well. SIQR model has also predicted that India will see the peak with 22,000 active cases during the last week of April followed by reduction in active cases. It may take complete July for India to get over with COVID-19.

## INTRODUCTION

Coronavirus disease 2019 (COVID-19) is originated from the Wuhan city of the Hubei province (China) in December 2019. This outbreak has caused global pandemic with more than 1.57 million positive cases and 92,191 deaths. India has witnessed its first positive case on 30^th^ January 2020, at present more than 6000 individuals are reported positive with 200 fatalities and 602 recoveries [1]. Many proactive steps are taken by the government to control the spread of disease, including lockdown, social awareness and identification of clusters of cases.

Mathematical equations are widely used to model the nature and impact of global pandemics in the society. The SIR model [2] is the classically adopted mathematical model to analyse and predict evolution of a disease. Its one of the variant SIQR [3] is considered to be the best modelling technique for COVID-19, where isolation of infectious plays an important role. This modelling approach to analyse the impact of this disease has been performed for few affected nations, including Brazil [4] and Italy [5].

Studies about the power-law growth [6] and the effect of lockdown on the spread of disease [7] have been reported for India. In this paper, we analysed the evolution of COVID-19 in India with the SIQR model. Parameters and indicators that quantify the growth and spread of disease in India are determined.

### MODELLING APPROACH

Susceptible Infected Quarantine Recovered (SIQR), a variant of classical SIR model appears to be particularly convenient for the modelling of COVID-19 [5]. This model considers two categories for infected individuals, one who gets quarantine and others who don’t (asymptotic or negligence). Susceptible in the models are the individuals who are at the risk of getting infected. Infected are those susceptible who gets affected by the virus; they may be asymptotic or have symptoms. Infected individuals who develop signs and gets isolated is considered as quarantine. Recovered are those infectious or quarantine individuals who recovered or died from the disease.

Rate equations of each part of the model are described using the following ODEs [3]: -

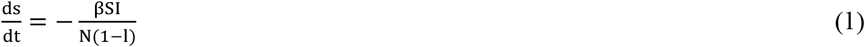

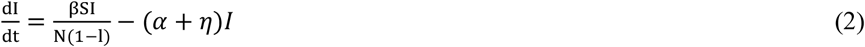

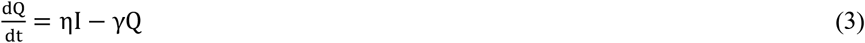

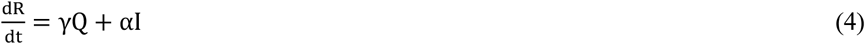

In the following equations, *β* denotes the rate of infection, *η* determines the rate at which new cases are detected from the infected population. *γ* is the rate at which quarantine are getting removed (recovered or died). *α* is the rate of removal of infectious individuals who are asymptotic (or for any reasons) and didn’t get quarantined (Fig. 1).

**Fig 1.**
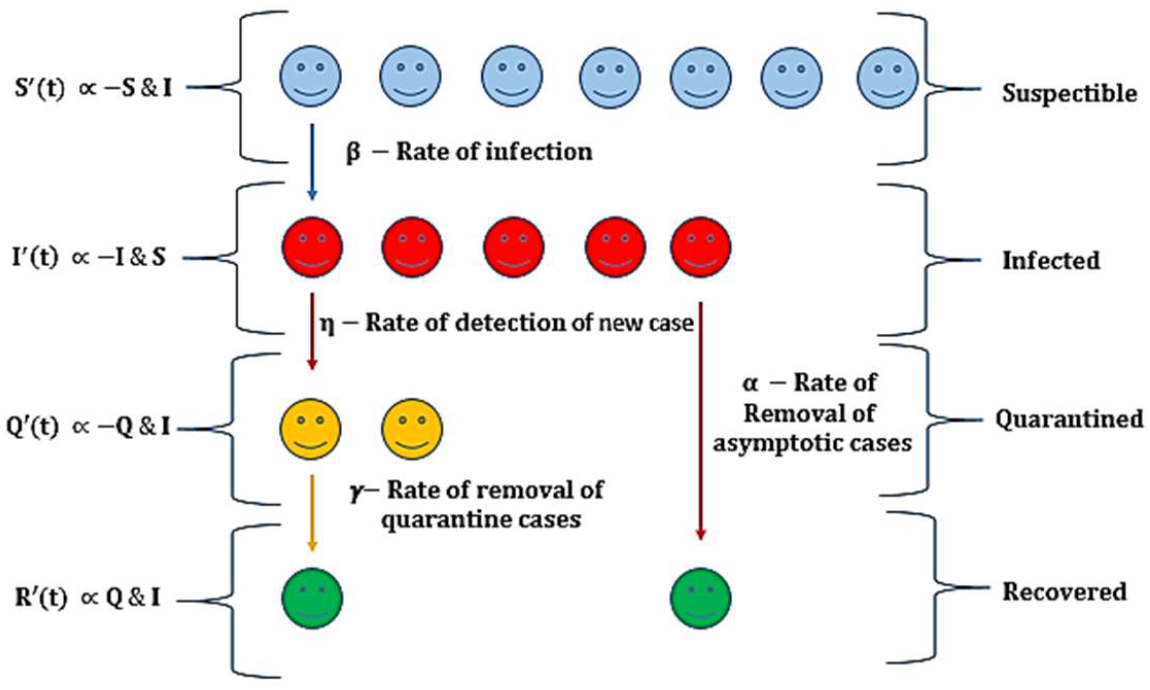
Pictorial representation of the dependence of each component of the SIQR model, where S stands for Susceptible, I for infected, Q for quarantined and R for recovered. Left side of the picture represents the dependence of the rate of change on S, I, Q, R. Four proportionality constants relating each parameter of the model is mentioned.

N is the size of the population. Complete lockdown of the country assures that the total population is not prone to the disease. A factor of (1-l) to N, with ‘l’ being the fraction of population following lockdown, is used to consider lockdown.

## RESULTS AND ANALYSIS

To estimate the parameters of Eq. (1-4), modified equations are fitted with the reported data of the country [1]. First positive case in the country was reported on 30^th^ January 2020 with steady growth till 1^st^ March, as the cases surge exponentially (Fig. 2). Considering the population not following lockdown to be susceptible, i.e. S ≈ N(1 − l) and approximating Eq. (2) to

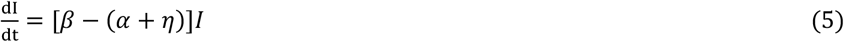

Integrating Eq. (5) to obtain

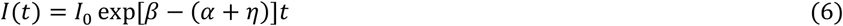

*I*_0_ is the number of infected individuals at the start of the disease in the country. Our study has considered the number of initial cases as *I*_0_ = 6, since cases were almost steady before that.

**Fig 2.**
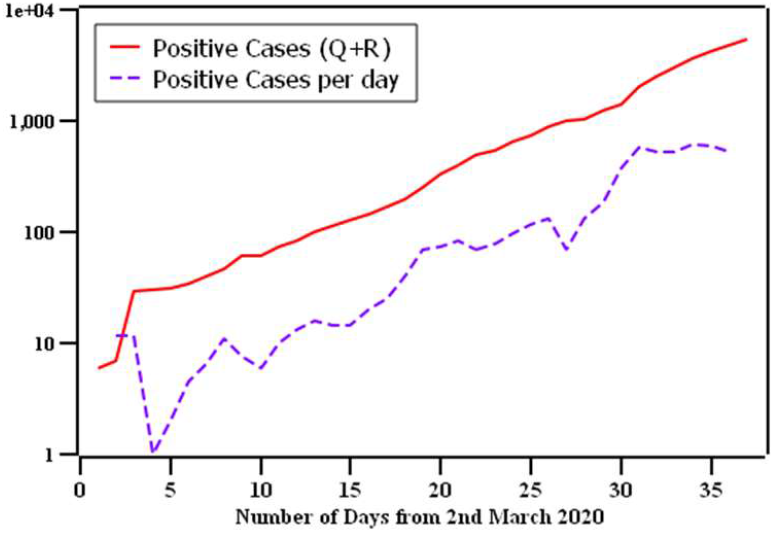
Semi-log (y) plot of the total confirmed positive cases and growth per day in India as of 7^th^ April 2020.

**Fig 2.**
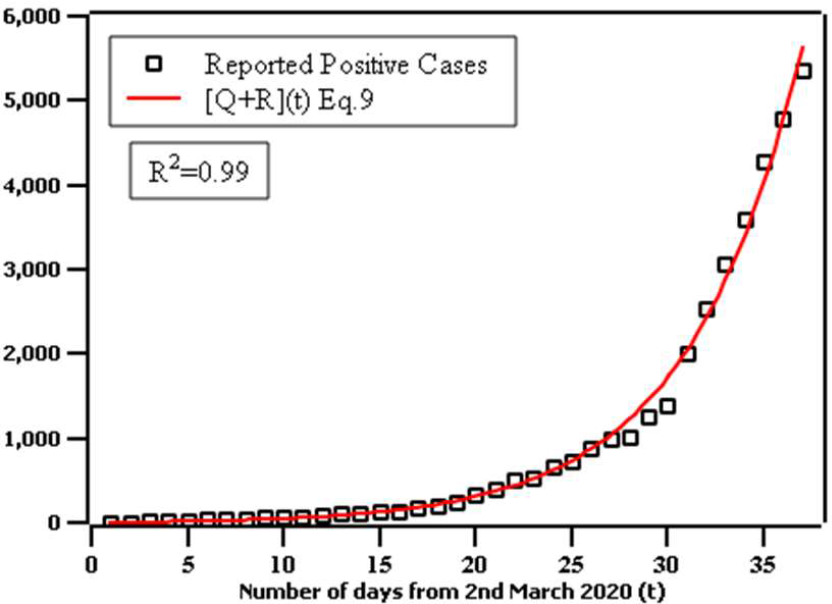
Least square fitting of Eq. (9) with the total positive reported cases from 2^nd^ March – 7^th^ April 2020. Fitted parameters are (*α* + *η*)*I*_0_ = 1.840 ± 0.201, *β* − (*α* + *η*) = 0.169 ± 0.004.

Adding Eq. (3) and Eq. (4) will give us the rate of change of the total confirmed positive cases [Q+R] in the country with the day (t).

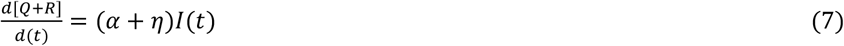

Putting *I*(*t*) from Eq. (6) to Eq. (7) will give us –

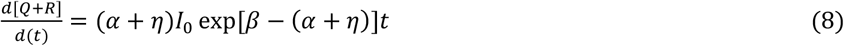

Integrating Eq. (8) over *t* –

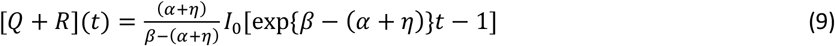

Above equation (Eq. (9)) is in the form of 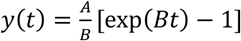 Where *y*(*t*) is the number of confirmed positive cases on a day (t). This form of equation is fitted to the total positive reported cases using least square fitting (Fig. 2) to give (*α* + *η*)*I*_0_ = 1.840 ± 0.201 and *β* − (*α* + *η*) = 0.169 ± 0.004. Fitted parameters give us *β* = 0.476 and *α* + *η* = 0.307.

Rate of detection of new cases from infectious individuals (*η*) can be calculated based on the report of the incubation period and fraction of infected population that gets quarantine. On average, it takes five days for an infectious individual to get symptoms [8]. If that individual gets quarantined on that day only, this rate can be written as *η* = 0.2 × µ, where µ is the fraction of the infected individuals that get quarantined. It has been reported that in Japan 50% of the population is asymptotic [9]. Considering the domination of young population in Indian age distribution [7] and low testing per million of the population [10]. It is assumed that 10% of the total infected population will get quarantined, i.e. µ is considered as 0.1. that leads to *η* = 0.2 × 0.1 = 0.02. It is not feasible to determine the rate at which quarantines are getting removed (*γ*), due to less number of recovered individuals in the country till 7^th^ April 2020. *γ* is assumed as 0.04 that means, on average, it took 25 days for a quarantined patient to get recover or die in line with the analysis presented in [5].

All these model parameters are used to determine indicators that quantify the transmissibility of disease in the country [4]. These indicators are defined as follows:

- Basic reproduction number (*R*_0_): This number is used to quantify the transmission ability of a disease. It is defined as the average number of individuals that can get infected from a single individual. It is formulated as follows:

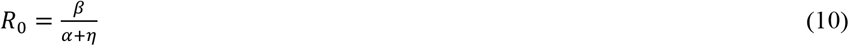
- Epidemic doubling time: Number of days required for a disease to double its infected population is epidemic doubling time. It is defined as:

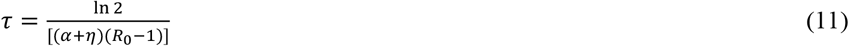
- Infected to Quarantine ratio: This ratio gives us the estimate of the population which are infected but not quarantine. They may be asymptotic or suffering from mild symptoms that get unnoticed.

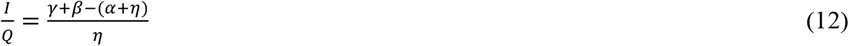

Values of the above-defined three indicators are mentioned in Table. 1 compared with the reported numbers from Brazil [4] and Italy [5]. India’s basic reproduction number is on the lower side as compared to Brazil and Italy, and this may be because of the strict lockdown and immediate isolation of the infected. Perhaps for the same reasons doubling time of the infected individuals is higher in India than that of Brazil and Italy. Whereas the ratio of actually infected to confirmed is more than ten times, highest among three. Probable reasons for this can be the presence of many asymptotic individuals in the population or the less number of testing. Temporal evolution of the number of active cases (Q) and the total infected population (I) is determined using estimated parameters. Numerical integration of Eq. (1-4) is performed with *N* = 1.3 × 10^9^ (population of India) with lockdown efficiency (l) determined by fitting active cases to the quarantine (Q) in Fig. 3.

**Fig 3.**
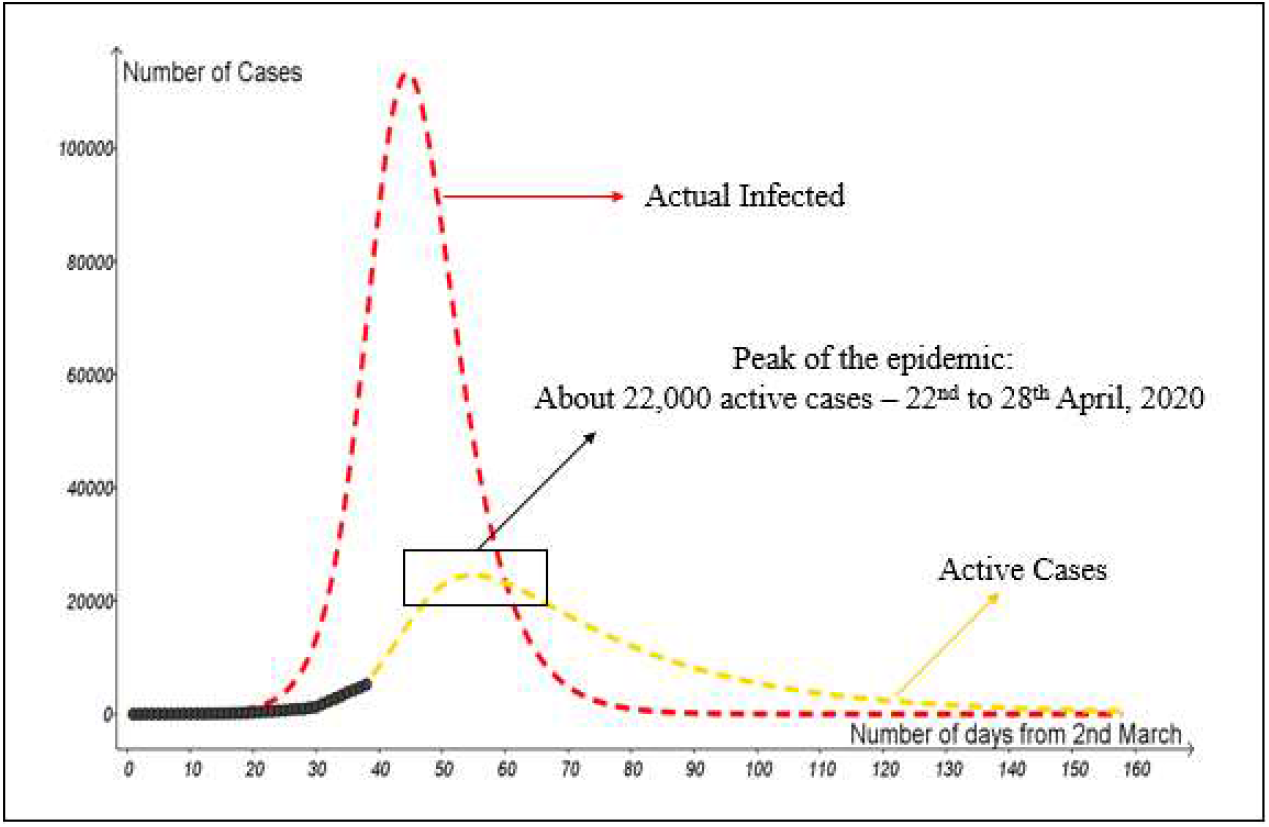
Temporal evolution of the number of active cases (Q) and actual infected (I) with the day (t) estimate using numerical integration of Eq. (1-4) with *α* = 0.287, *β* = 0.476, *γ* = 0.040, *η* = 0.020, *l* = 0.99942, *N* = 1.3 × 10^9^. Data of confirmed active cases are also marked (using black filled squares).

**Table 1.** Three different indicators calculated from the estimated parameters for India, Brazil and Italy.

| Indicator | India | Brazil | Italy |
| --- | --- | --- | --- |
| Basic reproduction number | 1.55 | 5.25 | 2.59 |
| Epidemic doubling time | 4.10 days | 2.72 days | 3.32 days |
| Infected to Quarantine Ratio | 10.45 | 9.83 | 3.7 |

Solution of ODEs estimated that around 7.54 × 10^5^ are susceptible to the disease. India may witness the peak of active cases in between 22^nd^ – 28^th^ April 2020 with about 22,000 total active cases and it may take complete July to finish this pandemic in India.

### CONCLUSION

Susceptible-Infectious-Quarantined-Recovered (SIQR) model is used in this paper to estimate the indicators of the spread of the COVID-19 in India. Following points sums up the key findings of this paper.

1. A single individual can infect 1.55 other individuals (i.e. *R*_0_ = 1.55) which is less than Brazil’s and Italy’s. It may be because of the strict lockdown or immediate isolation of the infected.
2. It takes 4.10 days in India to double the infected cases. Brazil and Italy take 2.72 and 3.32 days, respectively.
3. The ratio of actually infected to confirmed is more than ten times, higher than Brazil and Italy. Perhaps because of the presence of many asymptotic individuals or the less number of testings.
4. Our model also predicts India may witness maximum 22,000 active cases (total positive - recovered - died), somewhere in last week of April.
5. It may take complete July to finish this pandemic in India.

### DISCLAIMER

I am not an epidemiologist and analysis (or discussions) is based on an elementary mathematical modelling SIQR. This paper predicts the pattern and indicators based on the past data (2^nd^ March to 7^th^ April) and a few assumptions. Prediction of the peak of the active cases is made using the model; these dates and numbers must not be taken for any administrative decisions.

## Data Availability

All the data used in this paper is available on web

https://www.worldometers.info/coronavirus/

## ACKNOWLEDGEMENT

I wish to extend my special thanks to Prof. Manaswita Bose from IIT Bombay for regular discussion while writing and modelling.

## REFERENCES

1. https://www.worldometers.info/coronavirus/ [Last accessed on 10.04.2020].

2. Anderson, R.M. and May, R.M., Infectious Diseases of Humans. Oxford University Press. Oxford, 1991.

3. Hethcote, H., Zhien, M. and Shengbing, L., Effects of quarantine in six endemic models for infectious diseases. Mathematical biosciences, vol. 180, no. 1-2, pp. 141–160, 2002.

4. Crokidakis, N., Data analysis and modeling of the evolution of COVID-19 in Brazil, 2003.12150 (2020).

5. Pedersen, M.G. and Meneghini, M., Quantifying undetected COVID-19 cases and effects of containment measures in Italy, preprint 2020, available on-line at https://www.researchgate.net/publication/339915690.

6. Verma, M.K., Asad A. and Chatterjee, S., COVID-19 epidemic: Power law spread and flattening of the curve, preprint 2020, https://doi.org/10.1101/2020.04.02.20051680.

7. Singh, R. and Adhikari, R., Age-structured impact of social distancing on the COVID-19 epidemic in India, 2003.12055 (2020).

8. Li Q., Guan X., Wu P., Wang X, Zhou L., Tong Y., et al. Early transmission Dynamics in Wuhan, China, of Novel Coronavirus-Infected Pneumonia. N Engl J Med. 2020.

9. Japanese National Institute of Infectious Diseases, https://www.niid.go.jp/niid/en/2019-nCov-e/9417-covid-dp-fe-02.html. [Last accessed on 10.04.2020]

10. https://www.aa.com.tr/en/latest-on-coronavirus-outbreak/worldwide-covid-19-testing-ratio-per-country-million/1800124 [Last accessed on 10.04.2020]

